# Impact of Serum Sample Storage on the Stability of TRAIL, IP-10, and CRP: Implications for Biomarker Research Employing Biobanks

**DOI:** 10.1101/2025.01.22.25320787

**Authors:** Roy Kalfon, Ayelet Raz, Salim Halabi, Roy Navon, Tanya M. Gottlieb, Eran Eden, Mary Hainrichson

## Abstract

Serum biobanks are a valuable resource for proteomic screening, clinical validation and tool comparison studies. Protein integrity can be impacted by sample handling from blood draw until serum separation, by serum processing and freezing, and by storage time in the freezer. Since proteins are destabilized with different dynamics, it is critical to consider each biomarker’s behavior under the conditions relevant to a given biobank, before employing the biobank to study a biomarker panel. MeMed BV^®^ is a next generation host-based diagnostic test for discriminating between bacterial and viral infection that computationally integrates the circulating levels of three biomarkers: TNF-related apoptosis-inducing ligand (TRAIL), interferon γ-induced protein 10 (IP-10), and C-reactive protein (CRP). Here we report the impact of serum sample storage at −80°C on the stability of these three biomarkers. Notably, TRAIL levels decrease by approximately 30% after five years of storage at −80°C, while IP-10 levels decrease by only 17% and CRP levels do not decrease significantly. We conclude that long-term freezer storage differentially compromises the integrity of MeMed BV’s constituent biomarkers.

## 1. INTRODUCTION

Biobanks are becoming a pivotal resource for accelerating biomarker research.[1] The quality of biological samples within biobanks is a fundamental consideration. Various factors related to sample processing and storage can influence the biomolecular composition of the sample, potentially impacting the results of biomarker analysis.

Host-based diagnostics is an emerging field that leverages ‘omics’ with machine learning to create next generation biomarker tools that provide actionable insights into the body’s response to disease.[2] The biomarkers can be proteins, RNAs or other biomolecules, with the test output based on an algorithm. In the field of infectious diseases, various host-based diagnostics are in development or have been recently regulatory cleared to aid the clinician in deciding if there is an infection, if the infection is bacterial or viral, and how likely the patient is to deteriorate and develop sepsis. Many of these tools employ biobanks during their derivation and/or clinical validation,[3–7] and now biobanks are serving as a resource for performing comparative performance studies.

MeMed BV® is a next generation host-based diagnostic test for discriminating between bacterial and viral infection that computationally integrates the circulating levels of three biomarkers: TNF-related apoptosis-inducing ligand (TRAIL), interferon γ-induced protein 10 (IP-10), and C-reactive protein (CRP).[8] Its diagnostic accuracy in children and adults has been established in multiple clinical studies.[3,9–15] Since it is the first bacterial versus viral biomarker panel to be regulatory cleared, it is important to establish the biobank conditions that will enable it to serve as a comparator for similar tools currently under development. Here we report the impact of serum sample storage on the stability of these three biomarkers.

## 2. METHODS

### 2.1 Study design

Patients were recruited in accordance with International Conference on Harmonization - Good Clinical Practice (ICH-GCP) and in line with local regulatory requirements (IRB#: Carmel - 0100-18-CMC; Rambam - 0442-18-RMC). Before the venous blood draw, written consent was obtained from each participant using the Institutional Review Board (IRB)/Independent Ethics Committee (IEC)-approved Informed Consent Form (ICF).

### 2.2 Eligibility criteria

Main inclusion criteria: Age 18–80 (inclusive) at time of enrolment, clinical suspicion of acute bacterial or viral infection and current disease duration ≤ 7 days.

Main exclusion criteria: HIV, HBV, or HCV infection (self-declared or known from medical records), active inflammatory disease (e.g., IBD, SLE, JIA, RA, other vasculitis), suspicion of infectious gastroenteritis/colitis with diarrhea, major trauma and/or burns in the last 7 days and major surgery in the last 7 days.

### 2.3 Sample handling

Blood samples were collected into serum-separating tubes (SST) and handled according to the manufacturer’s instructions for use of the SST (Becton Dickinson, Plymouth, UK) and MeMed BV (MeMed, Israel), up until freezing (Figure 1). Briefly, samples were allowed to clot for 30 minutes prior to centrifugation for 10min, 1400g. Once centrifuged, the serum was aliquoted into 120 µL portions within 2 hours of blood collection and frozen at −80°C. The biomarker measurements were performed after the following time periods of storage at −80°C: t=0 (fresh sample), 24 hours, 3 months, 25 months and 5 years.

**Figure 1.**
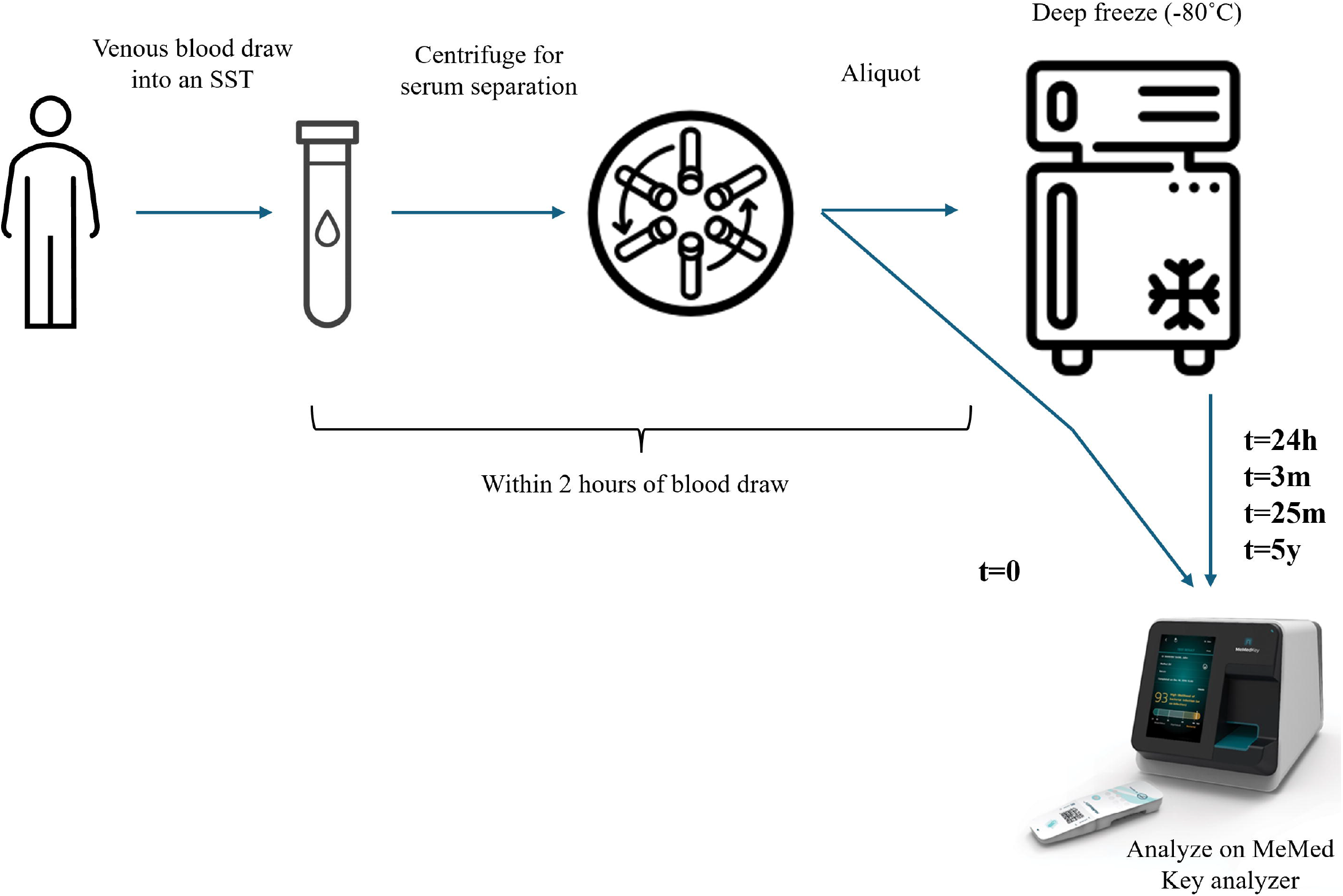
Schematic of sample handling.

### 2.4 TRAIL, IP-10 and CRP measurement

MeMed BV^®^ was run on the MeMed Key^®^ rapid platform. The MeMed BV test cartridges are single-use, multi-well containers that receive 100 µL of the patient’s serum sample, contain all the reagents and disposables necessary to conduct immunoassays and are the reservoirs for the waste. Upon insertion of the test cartridge into the MeMed Key analyzer, three independent immunoassays are conducted in parallel to measure TRAIL, IP-10 and CRP. Analytical validation of the system is described in Hainrichson et al, 2022.[16]

### 2.5 Statistical analysis

The sample size and analysis are in alignment with the “Measurement Procedure Comparison and Bias Estimation Using Patient Samples; Approved Guideline – Third Edition” (EP09c) issued by the CLSI.

Passing & Bablok regression was employed to evaluate the correlation between TRAIL, IP-10 and CRP concentrations at t=0 as compared to each storage duration.

## 3. RESULTS

TRAIL exhibited instability during frozen storage, with a gradual decrease in concentration over the storage period (Table 1 and Figure 2). After 2 years of storage at −80°C, the concentration of TRAIL was reduced by ~15%.

**Table 1.** Passing-Bablok regression values.

|  | t=24 hours | t=3 months | t=25 months | t=5 years |
| --- | --- | --- | --- | --- |
| **TRAIL** | 0.97x - 0.7 | 0.94x + 3.27 | 0.85x + 1.51 | 0.72x + 7.65 |
|  | R<sup>2</sup> = 0.99 | R<sup>2</sup> = 0.97 | R<sup>2</sup> = 0.91 | R<sup>2</sup> = 0.93 |
| **IP-10** | 0.99x - 1.62 | 1.07x - 2.4 | 0.93x - 13.21 | 0.83x - 3.77 |
|  | R<sup>2</sup> = 1.00 | R<sup>2</sup> = 0.99 | R<sup>2</sup> = 0.99 | R<sup>2</sup> = 0.99 |
| **CRP** | 0.97x - 0.03 | 1.00x - 0.0 | 0.99x + 1.04 | 1.04x - 1.69 |
|  | R<sup>2</sup> = 1.00 | R<sup>2</sup> = 0.97 | R<sup>2</sup> = 0.96 | R<sup>2</sup> = 0.96 |

**Figure 2.**
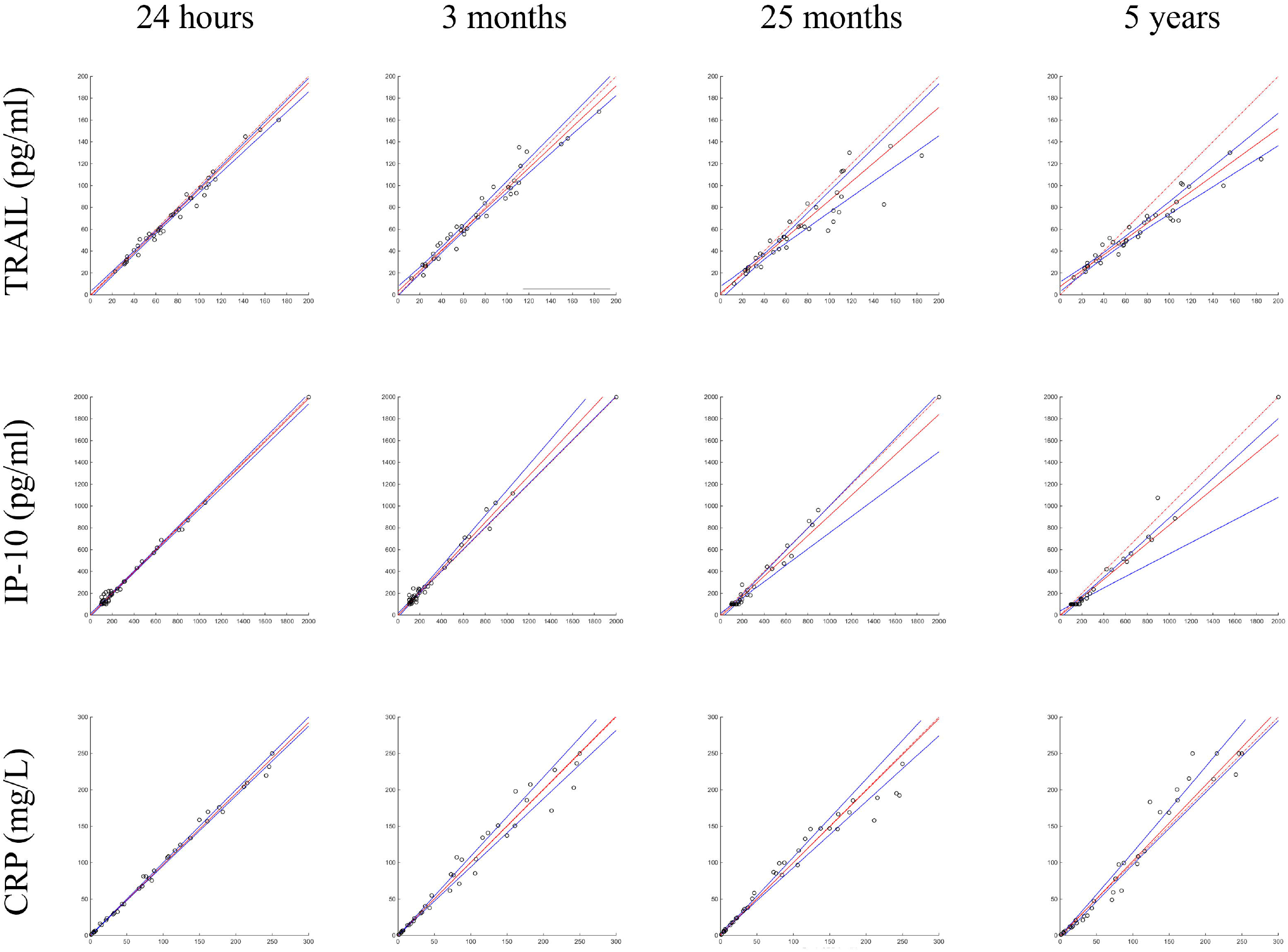
Passing-Bablok regression graphs. Analyte measurements were performed after storage at −80°C for the following time periods: 24 hours, 3 months, 25 months, and 5 years, compared to t=0 (fresh sample). Measurements were conducted using the MeMed Key. The x-axis represents biomarker concentration in fresh samples and the y-axis represents biomarker concentrations in frozen samples. The identity line is shown in red (dashed), with the regression line in solid red, and a 95% confidence interval in blue.

IP-10 demonstrated better stability, with only a small decrease in concentration after 2 years of storage at −80°C and a 17% decrease after 5 years. CRP demonstrated high stability throughout the 5-year storage period at −80°C.

## 4. DISCUSSION

This study establishes that TRAIL is unstable in serum stored for long periods of time at −80°C. In contrast, CRP was observed to be robust in serum stored at −80°C, in line with previous reports.[17,18] IP-10 exhibited intermediate stability during storage at −80°C.

Notably, 24 hours of storage at −80°C was not found to impact the stability of TRAIL, IP-10 and CRP in serum. This corroborates the manufacturer’s instruction for use that the MeMed BV test can be performed on serum samples that have undergone a single freeze-thaw.

Overall, we conclude that long-term freezer storage differentially compromises the integrity of MeMed BV’s constituent biomarkers. Therefore, we advise caution when considering measuring MeMed BV in biobanks that are over 2 years old.

## Data Availability

All data produced in the present study are available upon reasonable request to the authors.

## 5. ACKNOWLEDGEMENTS

We thank our colleagues for their input: Boris Lebedenko M.Sc. and Einav Simon Ph.D.

## 5.1 Author contributions

RK and RN took part in conceptualization, formal analysis, visualization, writing original draft, reviewing and editing of the final manuscript.

AR, SH, TMG, EE and MH took part in conceptualization, supervision, writing original draft, reviewing and editing of the final manuscript.

